# Associations between Chronic Stress, Resilience Resources, and Cardiovascular Health among Young Adults in Puerto Rico: the PR-OUTLOOK study

**DOI:** 10.64898/2026.03.18.26348758

**Authors:** Milagros C. Rosal, Sharina Person, Catarina I. Kiefe, Katherine L. Tucker, Cynthia M. Pérez

## Abstract

**Background:** Cardiovascular outcomes for young adults, particularly Latino individuals, have worsened, in contrast with trends for older persons. Stress and psychosocial resilience resources have been associated with cardiovascular health (CVH) among middle-aged and older adults, but these associations have not been characterized in young adults and Latino populations. We examined the association between chronic stress, resilience resources, and CVH in PR-OUTLOOK, a large community cohort study of 18-29 year olds residing in Puerto Rico.

**Methods:** Participants (n=2,676; 61.9% female) were assessed between September 2020 and March 2024. The American Heart Association Life’s Essential 8 (LE8), derived from surveys, laboratory assays, and physical examinations (range: 0-100, suboptimal CVH = <80) measured CVH. Surveys assessed chronic stress and resilience resources (optimism, religiosity, spirituality, and social support). Multivariable logistic regression, adjusting for age, sex, marital status, subjective social standing, and maternal education, examined associations between chronic stress and CVH, and the potential protective effect of resilience resources (moderation effect). With mediation analysis, using nonparametric bootstrap standard errors with 1,000 replications, we tested whether resilience factors were in the pathway of the stress-CVH association (mediation effect).

**Results:** High chronic stress was associated with suboptimal CVH (OR=1.46; 95% CI: 1.19, 1.80) and resilience factors did not moderate this association (all p > 0.05); however, optimism and social support mediated it, accounting for 26% and 10% of the association, respectively.

**Conclusions:** Chronic stress was associated with suboptimal CVH directly and indirectly through lower resilience resources. Longitudinal studies should better characterize these associations.

## Introduction

Current trends in cardiovascular disease (CVD) show steadily worsening CVD and CVD-related mortality among younger adults (<55 years) over time, in contrast with the observed declining trend among older individuals.^1,2^ This is particularly pronounced among young Latino adults, who have experienced increased stroke incidence,^3^ slower decline in stroke mortality, and acceleration in heart failure mortality;^4^ and have greater prevalence of traditional CVD risk factors,^5^ compared to non-Latino White individuals. Additional studies show that cardiovascular health (CVH), as measured by the American Heart Association (AHA) Life’s Essential 8 (LE8) for individuals across the lifespan,^6^ deteriorates rapidly during young adulthood,^7^ and CVD risk factors rise substantially from young adulthood to middle age, compared to any other life-course period.^8–10^ In the PR-OUTLOOK study, with a large community cohort of young adults (ages 18-29) residing in Puerto Rico, nearly two-thirds of participants experienced suboptimal CVH.^11^ As CVH predicts cardiovascular outcomes,^12^ a greater understanding of factors that may contribute to and protect against suboptimal CVH may reveal novel intervention targets for CVD prevention in this vulnerable population.

A body of literature has examined the influence of psychosocial factors, such as stress and resilience resources, on CVD progression and mortality among middle-aged and older people.^13–19^ However, associations between stress and CVH among young adults remain understudied,^20,21^ and few studies have focused on young Latinos. Young adulthood is characterized as a developmental period marked by multiple life changes, including moving away from home, the pursuit of higher education or professional training, entering the workforce, new financial responsibilities, and relationship and family-related stressors, among others. Whether chronic stressors preclude the ability of young Latino adults to achieve optimal CVH is unknown. Further, factors that may protect or amplify the influence of chronic stress on CVH among young adults remain to be identified. Individual and interpersonal resources associated with psychological resilience, defined as a person’s capacity to adapt positively upon encountering stress, adverse circumstances, or significant challenges,^22^ may mitigate or amplify associations between stress and health. Specifically, the Reserve Capacity Model and others have posed that resilience resources can attenuate the harmful effects of stress on health outcomes (e.g., stress-buffering or moderation hypothesis).^23,24^ Support for this hypothesis comes from a body of literature showing that, in the face of stress, resilience resources such as optimism, spirituality, religiosity, and social support are protective of CVH and other CVD outcomes.^25–27^ However, it has also been argued that chronic stress may deplete these resilience resources over time, limiting an individual’s ability to manage stress effectively.^28–30^ A decline in availability or access to resilience resources, in turn, would increase an individual’s vulnerability to the adverse health effects of stress (mediation hypothesis). This hypothesis, however, has received limited research attention.

Our study was driven by the assumption that a greater understanding of the associations between stress, resilience factors, and CVH among young adults can identify new intervention targets for prevention efforts, Therefore, we examined associations among chronic stress, resilience factors (i.e., optimism, spirituality, religiosity, and social support), and CVH among young Puerto Rican young adults in the PR-OUTLOOK study.^31^ We hypothesized that: 1) chronic stress would be associated with suboptimal CVH; 2) greater resilience resources would modify the adverse effect of chronic stress on CVH; and 3) the chronic stress-CVH association would be mediated by resilience resources such that greater stress would be associated with lower resources and suboptimal CVH.

## Methods

### Study design

We examined the association between chronic stress and CVH among young adults and the potential dual role of resilience factors, namely optimism, spirituality, religiosity, and social support, as moderators and mediators of such effect. Data are from the PR-OUTLOOK study, whose design and methods have been previously described.^31^ Briefly, cohort participants were young adults aged 18-29 residing in Puerto Rico. Participant eligibility criteria included (1) age 18 to 29 years, (2) self-identifies as Puerto Rican, born in Puerto Rico, and has at least one Puerto Rican-born parent; (3) resides in Puerto Rico; (4) has access to a phone; (5) has no history of cognitive, psychiatric, or physical conditions that would prevent the completion of study procedures; (6) is not on active military duty; and (7) has no immediate family member or partner participating in the study. Participants were recruited between September 2020 and March 2024 via public announcements on television, radio, social media, press releases, electronic and print advertisements, referrals, and community outreach activities. All participants consented in writing prior to participating in the study baseline assessment, which consisted of an online or telephone-administered survey and a clinical visit. The Institutional Review Board of the University of Puerto Rico Medical Sciences Campus approved the study (protocol number 2290033724R001).

### Measures

#### Cardiovascular Health (CVH)/Life’s Essential 8 (LE8)

CVH was measured using the AHA-recommended LE8 metric.^6^ Accordingly, each LE8 component (i.e., diet quality, physical activity, nicotine exposure, sleep health, body mass index (BMI), blood pressure, cholesterol, and glucose) was assessed as described below, each component metric was then calculated, and the eight component metrics were summed and divided by 8. The possible LE8 score range is 0-100. Scores ≥80 are considered ideal CVH, and scores <80 reflect suboptimal CVH.

Diet quality was assessed using the Modified Mediterranean Eating Pattern for Americans (MEPA) score,^32^ obtained from a validated food frequency questionnaire.^33,34^ Physical activity frequency, intensity, and duration were self-reported,^35^ as were nicotine exposure (use of combustible cigarettes or other nicotine-delivery systems) ^36^ and sleep duration (average time between sleep onset and waking during weekdays and weekends).^37^ Body mass index (BMI) was obtained from measured weight and height following standard protocols ^38^ (i.e., Tanita WB800-S Plus Digital Scale and weight rounded to the nearest tenth kilogram, and SECA 213 portable stadiometer with height rounded to the nearest centimeter), and BMI was calculated as weight in kilograms divided by height in meters squared. Systolic and diastolic blood pressures were measured three times at two-minute intervals after a 10-minute rest period (Omron HEM-907XL) and then averaged. Fasting total cholesterol, HDL cholesterol, triglycerides, and blood glucose were measured using the ARCHITECT Clinical Chemistry System. Non-HDL cholesterol was calculated as total cholesterol minus HDL cholesterol. Glycosylated hemoglobin (HbA1c) was assessed with a Tosoh Automated Glycohemoglobin Analyzer G8. Medication use was self-reported and verified with the drug containers provided by the participants. Assessments were conducted by rigorously trained staff.

#### Chronic stress

Chronic stress was assessed using the Chronic Burden scale.^39^ This tool asks about ongoing life stress across eight key domains in a person’s life, including personal and family health issues, job-related stress or unemployment, financial difficulties or instability, interpersonal conflict, alcohol or substance issues, and stress associated with caregiving. Participants are asked to indicate whether they have experienced stress in each of these domains, as well as the persistence (i.e., ≥ 6 months) and severity (i.e., not very stressful, moderately stressful, very stressful) of the stressor. A summary score was generated, reflecting the total number of stressors lasting at least six months that participants perceived as moderately or very stressful, with possible scores ranging from 0 to 8. The total score was dichotomized at the median due to its skewed distribution, with scores of 0–1 classified as low chronic stress and scores ≥2 classified as high chronic stress.

#### Resilience factors

Four resilience factors were assessed, selected based on existing literature and their relevance to Puerto Rican culture: optimism, spirituality, religiosity, and social support.^40^

Optimism was assessed using the Life Orientation Test-Revised (LOT-R),^41^ a widely used survey. This 10-item scale includes 3 items assessing dispositional optimism, or the degree to which a person has a general expectation that good things will happen in the future, and 3 questions assessing pessimism. Sample items include: “In difficult times, you usually expect the best,” and “You are always hopeful about your future.” Respondents are asked to indicate the degree to which they agree with each item using a 5-point Likert scale ranging from 1 (strongly disagree) to 5 (strongly agree). The optimism score is obtained by summing the optimism items and the reverse-scored pessimism items. Possible scores range from 0 to 24, with higher scores indicating greater optimism. The scale has good reliability and validity across a variety of populations, including cross-cultural samples, Spanish-speakers, and young adults. ^41–44^

Spirituality was assessed using the 16-item Daily Spiritual Experience Scale (DSES)^45,46^ designed to assess the frequency and nature of spiritual experiences in everyday life. The scale focuses on ordinary experiences of spirituality, rather than specific religious beliefs or practices, making it suitable for populations that do not adhere to a religious orientation. The tool assesses feelings of connection to a higher power, feeling guided by a higher power, a sense of peace or harmony, and compassion towards others. Sample items include “I feel God’s presence,” “I am spiritually touched by the beauty of creation,” and “I feel thankful for my blessings.” Respondents indicate the frequency of their experiences on a 6-point Likert scale 1 (many times a day) to 6 (never or almost never). Scores range from 16-96, with lower scores indicating more frequent spiritual experiences. The scale has shown good psychometric properties and has been extensively used with Spanish-speaking individuals and cross-cultural contexts. ^47^

Religiosity was assessed using a single question from the National Institute of Mental Health Collaborative Psychiatric Epidemiology Surveys.^48^ Respondents rate the importance of religion in their lives on a 4-point scale (4=“very important”, 3=“somewhat important”, 2=“not too important”, 1=“not important at all”). Possible scores range from 1 to 4, with lower scores indicating lower religiosity.

Social support from family and friends was assessed using the Social Support and Strain questionnaire.^49^ This scale assesses the perceived availability of support within a person’s interactions with their family and social network. Sample items include: “How much can you rely on them for help if you have a serious problem?”, and “How much do they really care about you?” Responses are rated on a 4-point Likert-type scale (1= “a lot”; 4= “not at all”). Items were reverse-coded so that higher scores reflect greater perceived support, yielding a total score of 2 to 8. The scale has shown good psychometric properties and has been used in previous large studies.^49,50^

#### Covariates

Covariates were selected a priori based on well-established associations with chronic stress and CVH. They included self-reported sex assigned at birth (male, female), age group (18–24 years, 25–29 years), marital status (unmarried, married/living together), maternal education (less than college, college or more), and subjective social standing (low: <5; high: >5).

### Statistical analysis

The analytic sample included 3,048 PR-OUTLOOK study participants (93.5% of enrolled participants). A total of 6 participants were excluded based on pregnancy status, and 366 were excluded due to missing data on any of the variables included in the analysis. Descriptive statistics (mean (SD) and frequency (percent)) were used to assess the sample distribution across study variables. The cross-sectional association between chronic stress and CVH was examined using multivariable logistic regression. The potential moderating effects of resilience resources were examined using additional multivariable logistic regression analyses incorporating interaction terms between chronic stress and each resilience factor, with statistical significance assessed using likelihood ratio tests. Next, associations between chronic stress and each resilience resource were evaluated using multivariable linear regression, and the associations between individual resilience resources and suboptimal CVH were assessed via logistic regression. Finally, mediation analysis was used to estimate the total, direct, and indirect effects of chronic stress on suboptimal CVH, with resilience resources examined as mediators.^51^ Analyses were conducted using logistic regression models. Mediation analyses were conducted under a no-interaction assumption, and additional sensitivity analyses included stress-resilience resource interaction terms. The natural direct effect (NDE) captured the effect of chronic stress on suboptimal CVH that did not operate through the mediator, whereas the natural indirect effect (NIE) represented the portion that operated through each resilience factor. The total effect reflected the combined direct and indirect pathways, and the proportion mediated (PM) was calculated as the ratio of the indirect to total effect. Standard errors and percentile-based bootstrap confidence intervals were estimated using 1,000 resamples. All analysis adjusted for the above-listed covariates and were conducted using Stata version 19.^52^

## Results

The sample was comprised of 2,676 young adults with an average age of 22.6 (SD=3.2) years (Table 1). Most participants were female, unmarried, had low subjective social status, and had mothers with at least some college education. More than one-third of participants reported high levels of stress, and the average LE8 score was 69.4 (11.9). Table 1 shows baseline sample characteristics for the entire sample and by chronic stress score (low vs. high) and CVH category (optimal vs. suboptimal). In bivariate analysis, individuals with high chronic stress were significantly more likely to be older (ages 25-29), female, and to report lower subjective social status, lower optimism, and lesser support from family and friends. Compared to those with ideal CVH, individuals with suboptimal CVH were significantly more likely to be male and married, and to report lower subjective social standing, lower maternal educational attainment, lower optimism, lower spirituality, and lesser social support from family and friends.

**Table 1.**
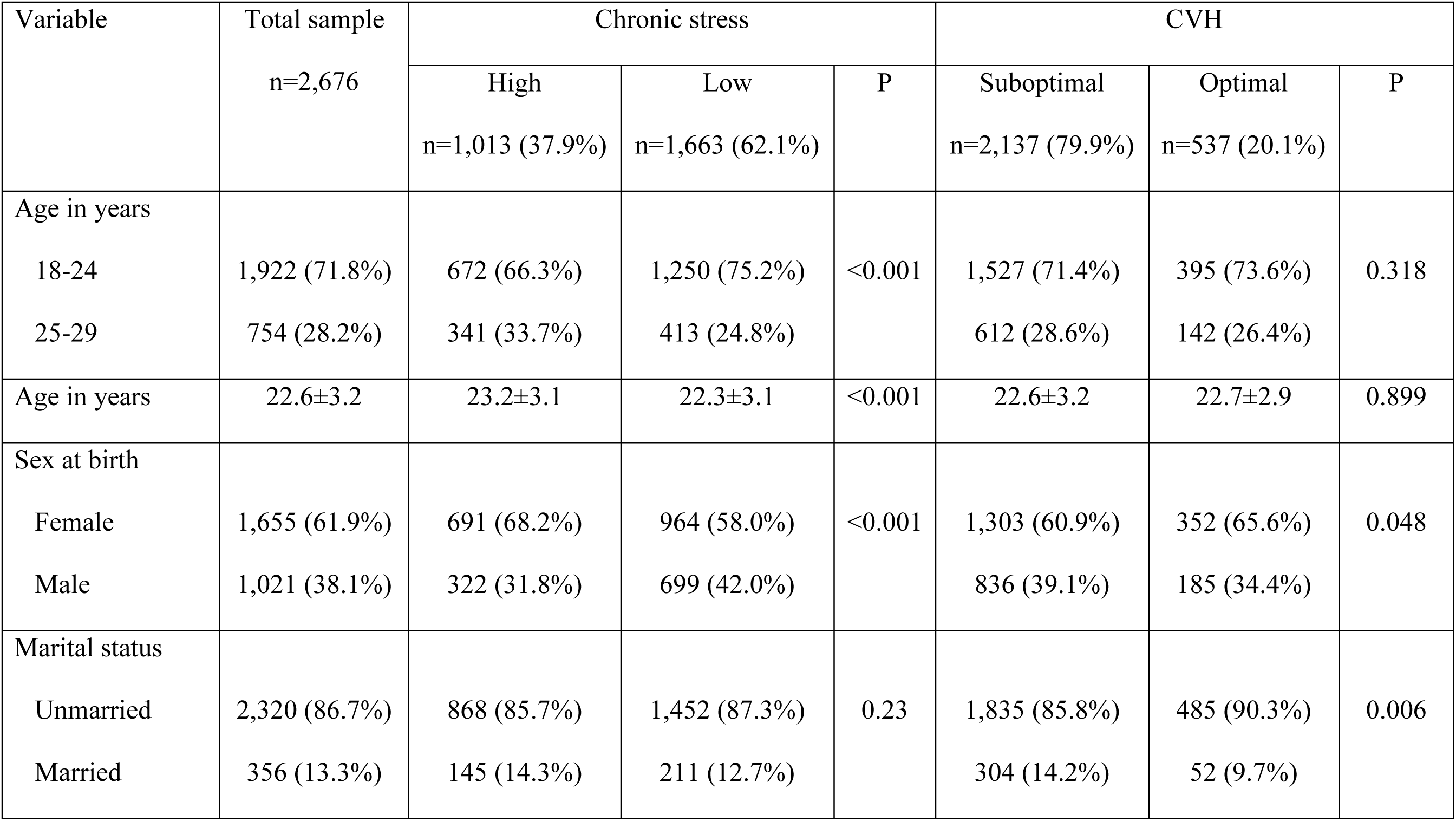
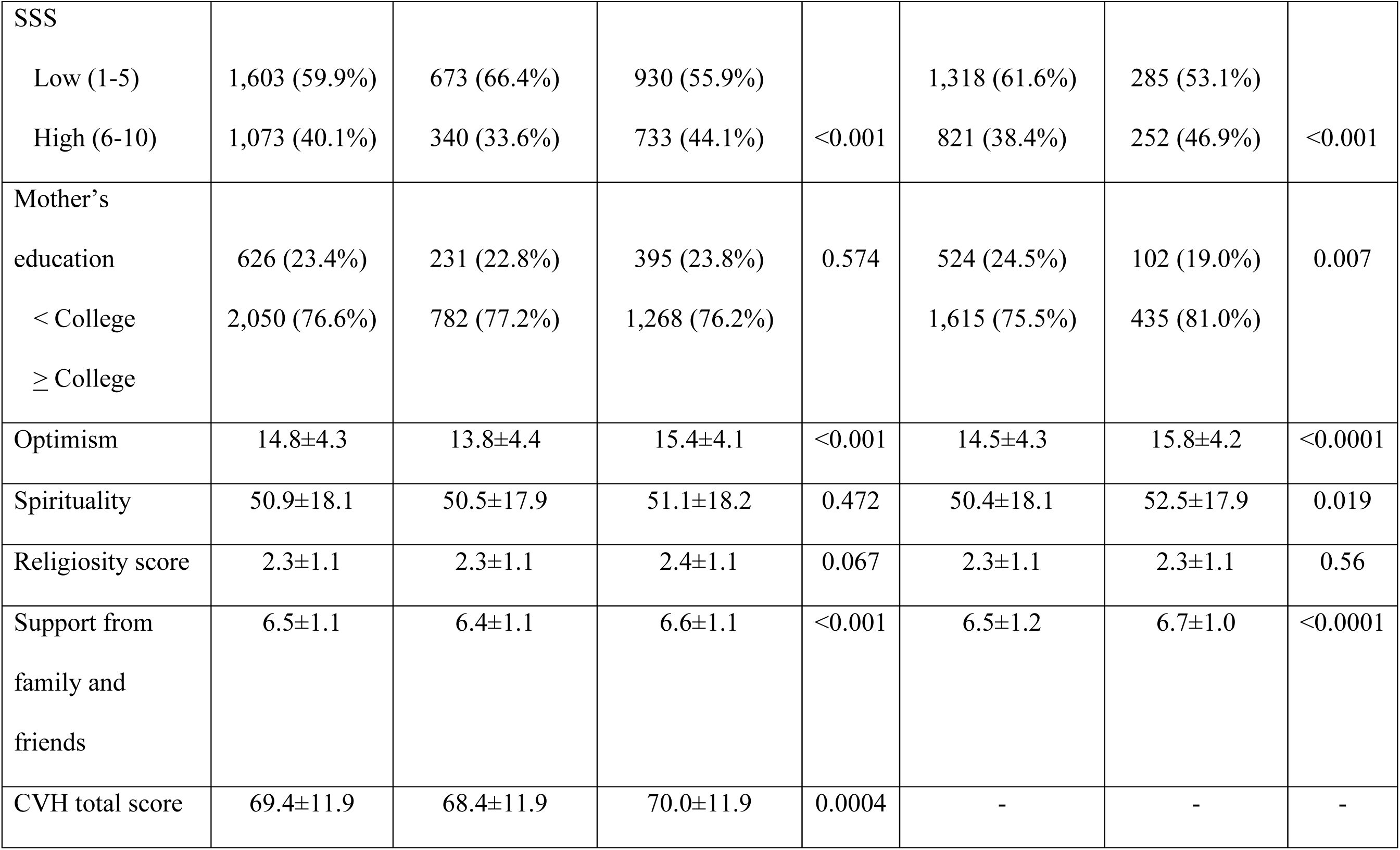
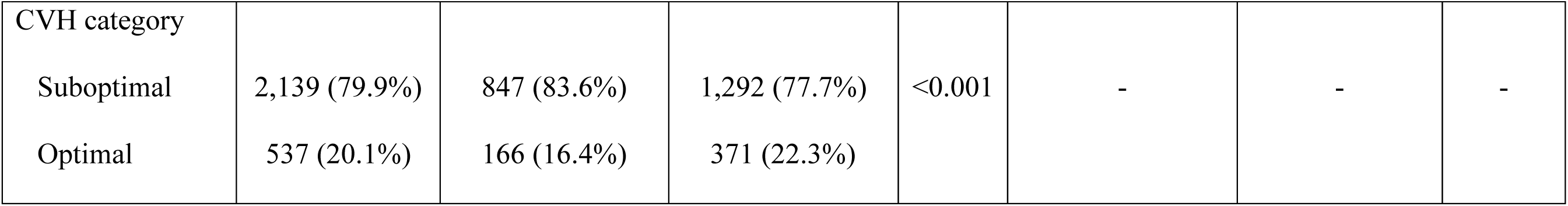
Baseline characteristics of participants overall and by chronic stress level: PR-OUTLOOK 2020-2024 (n=2,676).

### Association between chronic stress and CVH

In unadjusted analyses, experiencing high chronic stress (i.e., 2 or more chronic stressors) was associated with 47% higher odds of having suboptimal (vs. ideal) CVH, compared to experiencing low chronic stress (i.e., one or no chronic stressors) (OR=1.47 (95%CI: 1.20,1.80). This association remained statistically significant after adjustment for covariates, with those experiencing high chronic stress having 46% greater odds of experiencing suboptimal CVH, compared to those with low chronic stress (OR: 1.46 (95%CI:1.19, 1.80).

### Assessment of moderation by resilience factors

Analyses assessing whether individual resilience resources modified the association between chronic stress and CVH revealed no statistically significant interactions for spirituality, religiosity or support from family and friends (all interaction p-values > 0.50), and a trend for optimism (p = 0.065).

### Resilience resources as mediators of the chronic stress-CVH association

Linear regression analyses revealed that higher chronic stress was associated with lower levels of several resilience resources (Table 2). Specifically, high chronic stress was consistently associated with lower optimism and lower support from family and friends in unadjusted and adjusted models, and with lower religiosity in the adjusted model. However, there was no evidence of an association between chronic stress and spirituality in either model. Further, in unadjusted and adjusted logistic regression analysis examining resilience resources in relation to suboptimal CVH, higher optimism and greater support from family and friends were associated with lower odds of suboptimal CVH (Table 3). Spirituality was associated with lower odds of suboptimal CVH in unadjusted models, but this association was attenuated after adjustment. Religiosity was not significantly associated with suboptimal CVH in either model.

**Table 2.** Unadjusted and adjusted linear regression models examining associations between chronic stress and each resilience factor. PR-OUTLOOK Study, 2020-2024 (n=2,676)

|  | Model 1 |  | Model 2 |  |
| --- | --- | --- | --- | --- |
| | $\beta$ (95% CI) | P value | $\beta$ (95% CI) | P value |
| Chronic stress - Optimism | -1.65 (-1.98, -1.32) | <0.001 | -1.57 (-1.90, -1.24) | <0.001 |
| Chronic stress - Spirituality | -0.52 (-1.93, 0.89) | 0.472 | -1.15 (-2.56, 0.26) | 0.109 |
| Chronic stress - Religiosity | -0.08 (-0.17, 0.01) | 0.067 | -0.10 (-0.18, -0.01) | 0.034 |
| Chronic stress - Family and friends support | -0.24 (-0.33, -0.16) | <0.001 | -0.22 (-0.31, -0.13) | <0.001 |

\*Model 1 is unadjusted; Model 2 is adjusted for age, sex at birth, marital status, subjective social status, and maternal education.

**Table 3.**
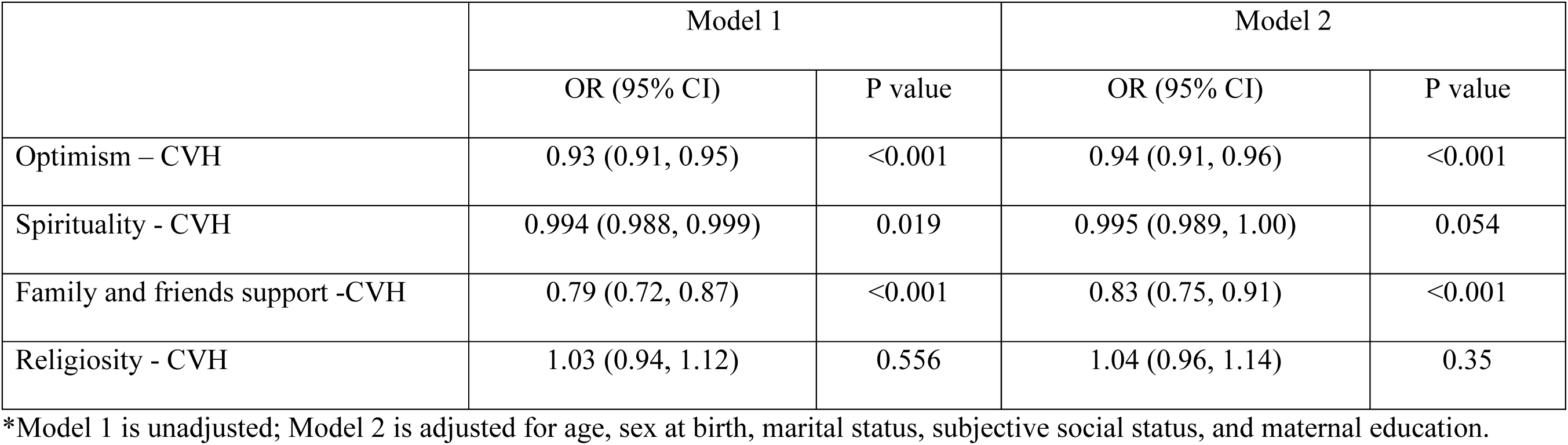
Unadjusted and adjusted logistic regression models examining associations between each resilience factor and suboptimal CVH. PR-OUTLOOK Study, 2020-2024 (n=2,676).

|  | Model 1 |  | Model 2 |  |
| --- | --- | --- | --- | --- |
|  | OR (95% CI) | P value | OR (95% CI) | P value |
| Optimism – CVH | 0.93 (0.91, 0.95) | <0.001 | 0.94 (0.91, 0.96) | <0.001 |
| Spirituality - CVH | 0.994 (0.988, 0.999) | 0.019 | 0.995 (0.989, 1.00) | 0.054 |
| Family and friends support -CVH | 0.79 (0.72, 0.87) | <0.001 | 0.83 (0.75, 0.91) | <0.001 |
| Religiosity - CVH | 1.03 (0.94, 1.12) | 0.556 | 1.04 (0.96, 1.14) | 0.35 |

\*Model 1 is unadjusted; Model 2 is adjusted for age, sex at birth, marital status, subjective social status, and maternal education.

Based on these analyses, separate single-mediator models were estimated for optimism and for family and friends’ support to examine their potential mediating roles in the association between chronic stress and suboptimal CVH, under the assumption of no interaction between stress and resilience resources (consistent with the findings above). Both optimism and support from family and friends partially mediated this association, accounting for 26.0% and 10.0% of the estimated total effect, respectively, after adjustment for covariates (Figure 1). Sensitivity analysis that included stress-resilience resource interaction terms yielded similar conclusions, though mediated proportions were attenuated (13.8% and 10.2% of the estimated total effect, respectively, after adjustment for covariates).

**Figure 1.**
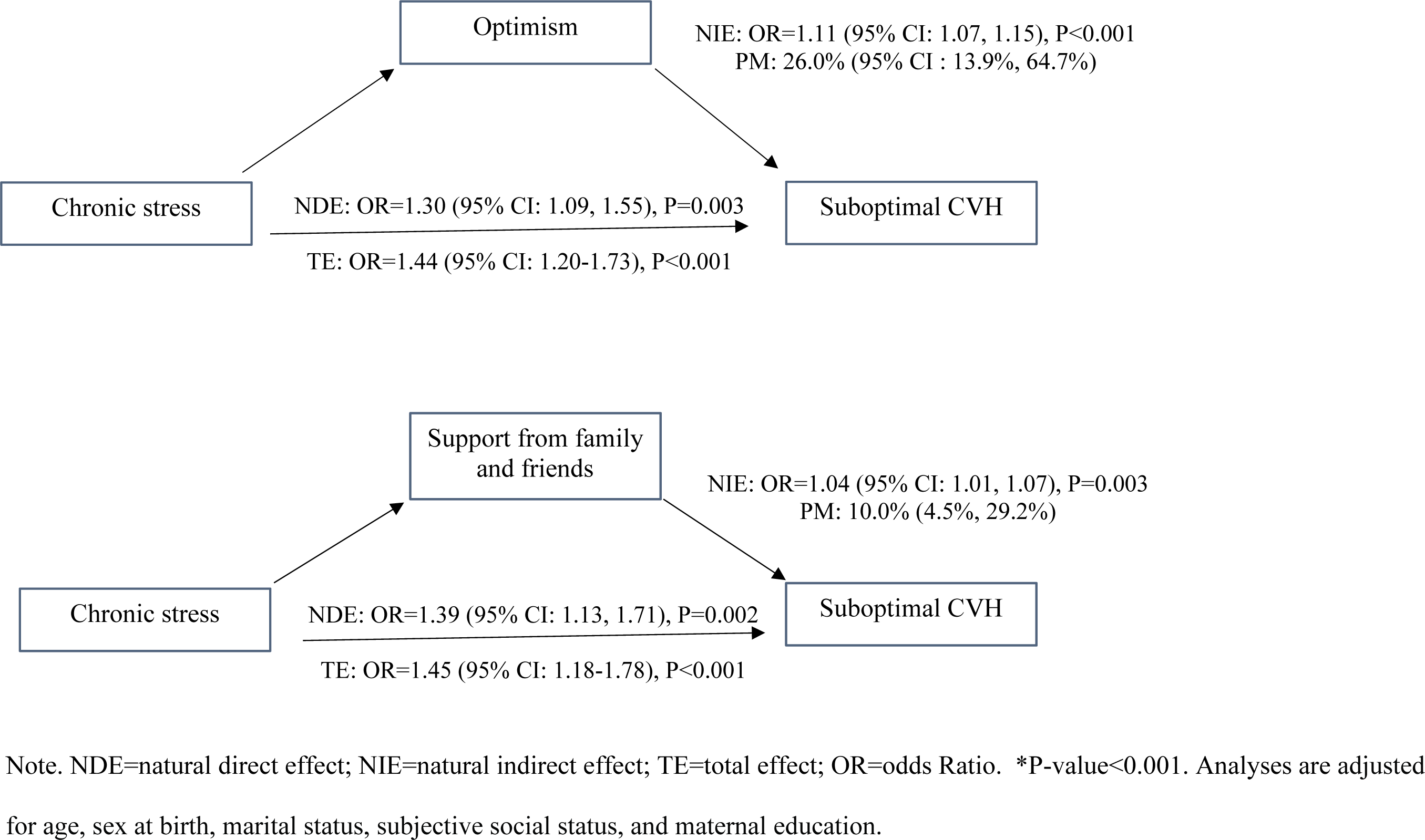
Mediation analysis of the associations between high chronic stress and suboptimal CVH, with optimism and family and friends’ support examined as separate potential mediators. PR-OUTLOOK Study, 2020-2024 (n=2,676).

## Discussion

This study examined the association between chronic stress, resilience resources (i.e., optimism, spirituality, religiosity, and social support), and CVH among a large community cohort of young adults in Puerto Rico. Findings confirmed two of our three hypotheses. Greater chronic stress was associated with suboptimal (vs. ideal) CVH, and this association was partially mediated by optimism and family and friends’ support. Greater resilience resources did not modify the association between chronic stress and CVH, as we had hypothesized. This study contributes to the growing interest in understanding how stress and resilience factors influence health and CVH, in particular. This is one of few studies that have examined these associations among young adults and within a cohort of Puerto Ricans. US Latinos experience a considerable burden of CVD risk factors compared to other population groups, and Puerto Ricans show particularly high risk factor prevalences compared to other Latino subgroups.^53–55^ Thus, improving our understanding of factors that may increase or protect from risk among young Puerto Ricans is essential to optimizing CVH and preventing the future burden of CVD in this population.

A large proportion of young adults in the cohort (37.9%) experienced high chronic stress and, consistent with our first hypothesis, the experience of high chronic stress resulted in a significant increase in the odds of having suboptimal CVH. While this was a cross-sectional analysis and we are, thus, unable to determine causality, this finding is consistent with evidence suggesting that chronic stress responses may promote adverse behavioral (i.e., smoking^56^and unhealthy dietary habits^57^) as well as physiological (immune, vascular, and metabolic) responses that increase cardiometabolic risk (i.e., hypertension,^58^abdominal adiposity,^59^and diabetes^60^), among a variety of populations including Latino individuals.^61–63^ This study provides further evidence in support of an association between chronic stress and CVH among young Puerto Rican adults, a vulnerable population.

While previous studies have shown that the availability of resilience resources is associated with better CVH, less is known about how such resources may influence health in the face of chronic stress. Our study sought to understand whether and how resilience resources might operate within the context of the relationship between chronic stress and CVH in this cohort of young adults. The Reserve Capacity model^23^ posits that the use of resilience resources is critical for managing stress and attenuating its negative impact on health. In this framework, psychosocial resources such as optimism and social support are expected to buffer the adverse effects of chronic stress on cardiovascular outcomes. In this study, however, we did not observe strong evidence of stress-buffering moderation. The interaction between chronic stress and optimism did not reach statistical significance, and no evidence of interaction was observed for spirituality, religiosity, or social support. Although detecting interaction effects typically requires greater statistical power than detecting main effects, the overall findings are consistent with resilience resources not meaningfully altering the association between chronic stress and CVH in this sample.

Our findings provide evidence of partial mediating effects for optimism and family and friends’ support, in line with our third hypothesis. Specifically, greater chronic stress was associated with suboptimal CVH in part because chronic stress was associated with reduced optimism and family and friends support which, in turn, was associated with suboptimal CVH. We are not aware of previous studies that examined resilience resources as mediators of the association between stress and CVH, and relatively few have examined them as mediators of other health outcomes. For example, a study of a large national sample of middle-aged and older adults reported that optimism did not modify the association between lifespan stress exposure and blood pressure reactivity, but lower optimism partially accounted for the stress-blood pressure reactivity association, ^64^ and another study conducted in Turkey reported that COVID-19-related stress led to lower optimism among young adults, and lower optimism mediated an association between stress and mental health.^65^ Previous studies have reported significant associations between exposure to adversity and reduced resilience resources,^66,67^ and others provide evidence of inverse associations between optimism and CVH among young and middle-aged individuals.^68–70^ Our findings contribute to existing literature by examining optimism as a mediator of the chronic stress – CVH association. It is important to note that optimism is a trait and thus considered a relatively stable personality characteristic, and a large body of research has focused on its potential to buffer the negative impact of stress on mental and physical health (i.e., moderation effects). However, our findings suggest that the experience of chronic stress among young adults may lead to a reduction in the level of optimism, and this effect may, in turn, contribute to the deterioration of CVH. This hypothesis will need to be examined in future longitudinal studies.

Our findings also showed that support from family and friends partially mediated the relationship between chronic stress and CVH. With some exceptions,^71,72^ a number of studies provide evidence concluding that individuals with better social relationships are more likely to achieve or maintain ideal CVH, as measured by the earlier CVH metric (LS7).^73,74^ However, few studies have examined a mediating role for social support on the link between chronic stress and CVH among adults of any age. A large study of young adults in the US found that social support mediated the link between perceived stress and physical and mental well-being such that, under higher chronic stress, young adults reported less available support, and this reduction in social support partially explained the association between high stress and poorer well-being. The authors argued that chronic stress may erode or strain relational resources (e.g., less time/energy to invest in relationships, more conflict), reducing access to support and amplifying the experience of stress.^28^ Another study found that early-life stress (i.e., child neglect and maltreatment) was associated with difficulty accessing and benefiting from social support resources in adulthood, and social support mediated an association between stress and mental health, and also moderated this association among men.^29^ Our study adds to emerging evidence of a mediation effect for social support in a sample of young adults.

Our study showed that, while statistically significant, the examined associations were small. However, if stress has the potential to undermine optimism and support from important relationships in the lives of young adults, which may effectively reduce coping resources and intensify the impact of stress on health among young people, this may have an important cumulative impact over the long term. Again, longitudinal studies that examine these associations over time are needed to determine their stability and examine causality.

Findings showed no evidence of mediation effects by religiosity or spirituality. We are not aware of studies that have examined religiosity or spirituality as mediators of the stress-CVH association. However, in contrast with our hypothesis that chronic stress may deplete resilience resources, recent studies suggest that stress may not have a uniform impact on religiosity or spirituality. A recent study showed that stress may amplify religious beliefs among strong believers and deepen skepticism among non-believers,^75^ and similar findings have been reported for spirituality,^76,77^ which could partially explain our findings.

This study has several limitations. The cross-sectional and observational nature of the study precludes causal conclusions. Indeed, associations between stress and resilience resources may be reverse or bidirectional. Second, although the study focuses on a unique population that has been underrepresented in research, findings may not generalize to individuals of other ages or ethnicities. Third, the effect sizes were small. However, the findings were statistically significant, and even small effects can be meaningful at the population level or when considering the effect of these associations across the lifespan.^69^ Additional considerations include potential self-selection and self-report biases, including recall and willingness to report sensitive information, such as stressful experiences.

This study also has several important strengths. This is the first study to examine stress, resilience factors, and CVH among an island-based cohort of young Puerto Rican adults. In addition to providing evidence of the harmful effect of chronic stress on CVH, the study contributes to the conceptual understanding of how stress and resilience may operate to influence health, with potential practical applications for designing preventive interventions. Most studies have examined the direct and buffering/moderating effect of resilience resources. Our findings help address a gap in the literature by examining the mediating role of resilience factors in the chronic stress-CVH link. We used the most recent measure of CVH, Life’s Essential 8, with all components measured using standard surveys and clinical and lab assessments. This is one of the few studies examining psychosocial factors associated with this metric among young adults and Latinos.

Improving cardiovascular health across all population groups is a Healthy People 2030 priority.^78^ Study findings show that many young adults in Puerto Rico experience chronic stress and suboptimal CVH, and may benefit from stress reduction interventions. Our data suggest that such interventions may be strengthened by including components that preserve resilience resources such as optimism and social support. While there is emerging evidence of a potential benefit of stress reduction interventions for CVD prevention,^79^ clinical trials of such interventions are needed to determine whether and what approaches work best for preserving and optimizing CVH, and reducing the future burden of CVD, in vulnerable populations of young adults, such as Puerto Ricans.

Concluding, this study provides evidence that chronic exposure to stress during young adulthood is associated with suboptimal CVH directly and indirectly through lower optimism and support from family and friends. As psychological stress is an unavoidable part of life and may be especially prevalent among young adults, there is a need for longitudinal studies to understand the mechanistic links between stress and CVH, and to test the impact of stress reduction interventions, enhanced to also target optimism and social support, on CVH in this population. Individuals with favorable CVH have substantially lower risk for CVD and mortality,^80^ thus preserving CVH in young adulthood has the potential to reduce CVD burden among Puerto Ricans.

## Data Availability

Data are available upon request.

## Acknowledgments

MCR, CMP, and CK conceptualized and designed the study. CMP analyzed the data. SP provided guidance for data analysis and interpretation. MCR drafted the manuscript. All authors reviewed, contributed to revisions, and approved the final version of the manuscript. The corresponding author confirms that all the listed authors meet the authorship criteria and that no eligible contributors have been excluded.

## Non-standard Abbreviations and Acronyms

CVH: Cardiovascular health
CVD: Cardiovascular disease
CI: Confidence interval
LE8: Life’s Essential 8

